# Modeling and predicting the spread of COVID-19 in Lebanon: A Bayesian perspective

**DOI:** 10.1101/2020.04.29.20082263

**Authors:** Samer A Kharroubi

## Abstract

In this article, we investigate the problem of modelling the trend of the current Coronavirus disease 2019 pandemic in Lebanon along time. Two different models were developed using Bayesian Markov chain Monte Carlo simulation methods. The models fitted included Poisson autoregressive as a function of a short-term dependence only and Poisson autoregressive as a function of both a short-term dependence and a long-term dependence. The two models are compared in terms of their predictive ability using root mean squared error and deviance information criterion. The Poisson autoregressive model that allows to capture both short and long term memory effects performs best under all criterions. The use of such a model can greatly improve the estimation of number of new infections, and can indicate whether disease has an upward/downward trend, and where about every country is on that trend, so that containment measures can be applied and/or relaxed.

## Introduction

As the Coronavirus Disease 2019 (COVID-19) pandemic progresses, countries around the world, including Lebanon, are increasingly implementing a range of responses that are intended to help preventing the transmission of this disease. Until a COVID-19 vaccine becomes available, strict measures from closing schools and universities to locking down entire cities and countries were inforced to suppress the virus transmission, thereby, slowing down the growth rate of cases and rapidly reducing case incidence.

Alternatively, structured mathematical and statistical techniques can be potentially powerful tools in the fight against the COVID-19 pandemic. These techniques allow the COVID-19 transmission to be modelled, so the resulting models can be used to predict and explain COVID-19 infections. This may be of great usefulness for health care decision-makers, as it gives them the time to intervene on the local public health systems, thereby take the appropriate actions to contain the spreading of the virus to the degree possible. Since the outbreak of the pandemic, there has been a scramble to use and explore various statistical techniques, and other data analytic tools, for these purposes.

Advancement in statistical modelling, such as Bayesian inference methods facilitate the analysis of contagion occurrence through time. It is well documented that infectious diseases grow exponentially and are usually driven by the basic reproduction number *R* (see for example [1]) for a given population. The value of R can be calculated by the ratio between the new occurrences arising in consecutive days: a short-term dependence. However, incubation time varies among individuals and incidence and so measurement may not be uniform across different populations. This implies that a long-term dependence should then be induced [2].

The aim of the present paper is to model and predict the number of COVID-19 infections in Lebanon using Bayesian methods. We present two different models of increasing complexity using Bayesian Markov chain Monte Carlo (MCMC) simulation methods. These models include Poisson autoregressive as a function of a short-term dependence and Poisson autoregressive as a function of both a short-term dependence and a long-term dependence.

## Methods

### Data source

The Ministry of Public Health has started to release a daily bulletin about COVID-19 infections in Lebanon since 23 February 2020. Data are available from the website of the Ministry of Public Health (MoPH) [3] and worldometer website [4]. The overall temporal distribution of daily counts of COVID-19 cases (blue line) is presented in Figure 1. The data covers the period from February 23 to April 18, 2020. The plot indicates that COVID-19 contagion in Lebanon has achieved a complete cycle. Specifically, Figure 1 shows an upward trend until a peak is reached on March 20: after this date a decreasing trend is observed.

**Figure 1.**
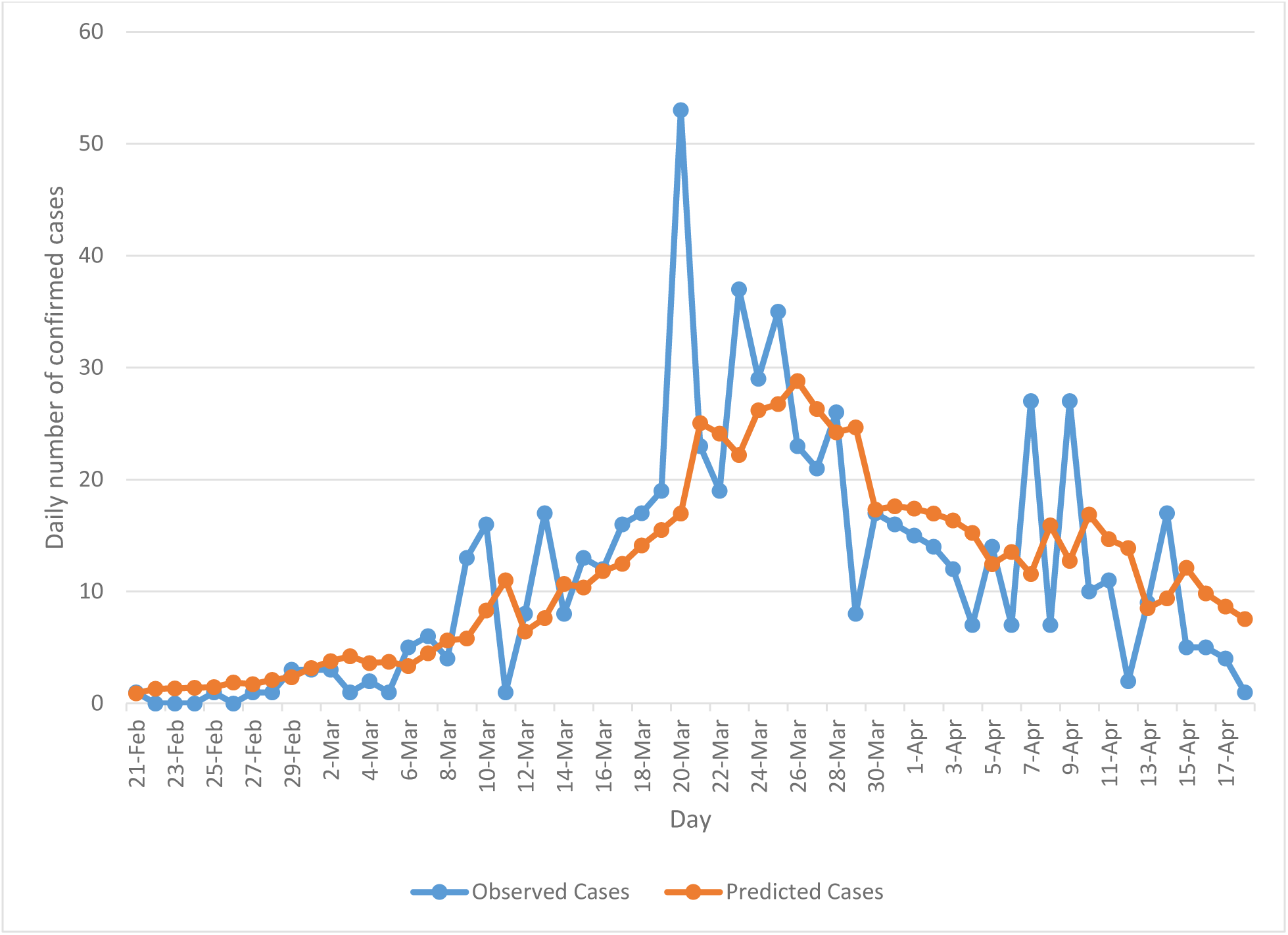
Actual (blue line) and predicted (orange line) cases based on the Poisson autoregressive model (Model 2).

### Model development

Two different models of were fitted to the data as follows:

1. Poisson autoregressive as a function of a short-term dependence only
2. Poisson autoregressive as a function of both a short-term dependence and a long-term dependence

Following Agosto and Giudici [2], the number of new cases *y_t_* reported at time (day) *t* is assumed to follow a Poisson distribution i.e.

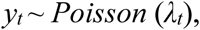

with a log-linear autoregressive intensity specification, as follows:

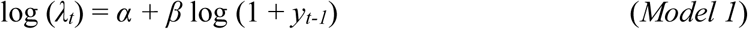

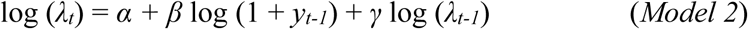

In each model, the inclusion of 1 in log (1 + *y_t-1_*) allows to address the problem generated by zero values, *α* represents the intercept term and *β* expresses the short-term dependence of the expected number of cases reported at time *t*, *λ_t_*, on these observed in the previous day (time *t* – 1). The *γ* component in model 2 corresponds to a trend component and, more specifically, it represents the long-term dependence of *λ_t_* on all past counts of the observed process. Note that the use of a log-linear autoregressive intensity specification, rather than linear, allows for negative dependence. See Agosto and Giudici [2] for further details on this.

### Model estimation

Both models were implemented from a Bayesian perspective using Gibbs sampling MCMC simulation methods using WinBUGS software [5]. The relevant code to undertake the Bayesian models is available on request. For every model, an initial 10,000 iterations were run as a “burn-in” to reach convergence [6]. This was evaluated by testing the convergence statistic for two parallel MCMC chains with different initial values. Following that, an additional 50,000 iterations were run for parameters estimation purposes. To this end, the prior distributions for all the regression parameters were specified as

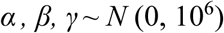

### Model validation

The two models were compared in terms of their predictive performance using root mean squared error (RMSE) and deviance information criterion (DIC) [5].

## Results

Table 1 shows the estimated autoregressive coefficients for both models together with their associated 95% credible intervals. We notice that all coefficients had the expected positive sign as well as their credible intervals excluding zero, indicating the presence of a short-term dependence for model 1 and both a short-term dependence and a long-term trend for model 2. A testing of the models’ performance is also shown in Table 1, where model 2 was found to perform best by scoring the best RMSE and DIC with 7.68 and 444.7, respectively, in comparison to model 1 (RMSE = 8.56, DIC = 517.5).

**Table 1.**
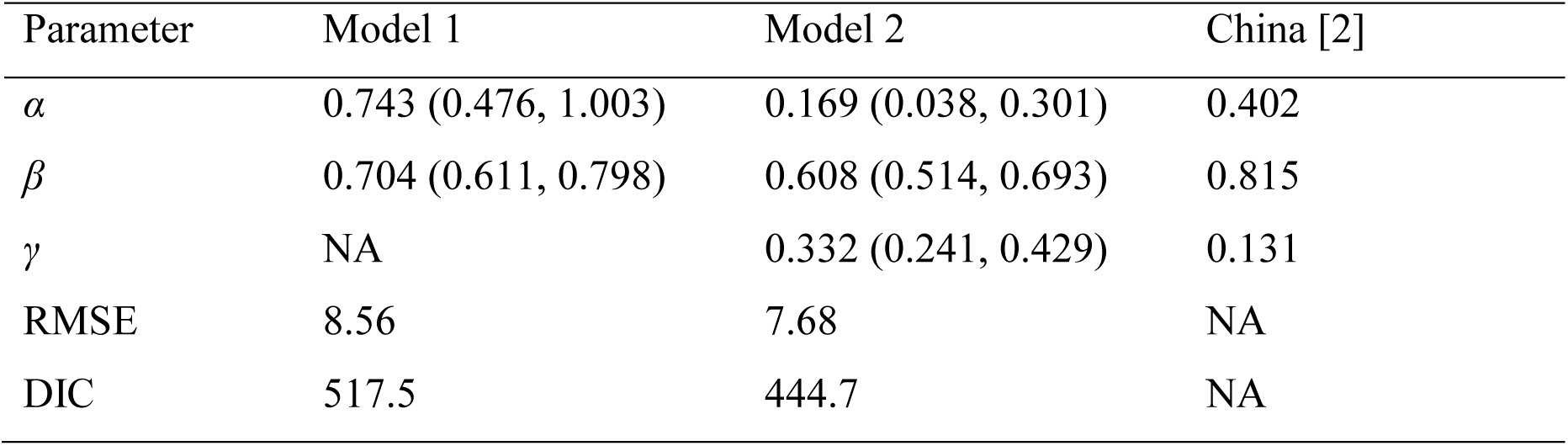
Model coefficients and model performance.

Model 2 has been tested in terms of its predictive ability where the resulting predicted mean occurrences (orange line) have been plotted along with the actual occurrences (blue line) in Figure 1. As can be seen from the plot, the Poisson autoregressive model as a function of both a short-term dependence and a long-term dependence predicts the data quite well. With aim of better interpreting the short-term time series and long-term time series of model 2, we notice from Table 1 that the estimated *γ* parameter is lower than *β*, confirming that a downward trend data is accumulated. Additionally, we splitted the Lebanese data into two-time periods and we repeatedly fitted the model for each data set. More specifically, we first fitted the model on the data, which covers the period from February 23 to March 20, 2020 and the analysis revealed that *γ* was larger than *β*, confirming the presence of an upward trend (the *γ* component) which absorbs the short-term component. After this date, the estimated *γ* parameter became lower, so a downward trend data is accumulated.

Agosto and Giudici [2] drew a similar conclusion from their analysis of the Chinese data, which covers the period from January 20 to March 15, 2020. Their estimated *β* and *γ* parameters (final column of Table 1) revealed that the contagion cycle was in a downward trend (*γ* < *β*). On the other side, their analysis for South Korea revealed non-significant estimate for the estimated *β* parameter, confirming absence of a trend effect on the daily cases. However, for Italy, their results showed that the estimated *β* parameter was smaller than *γ*, suggesting that the trend of the contagion has not peaked yet.

## Discussion

In this article, we have analyzed, by modelling, the trend of the current Coronavirus disease 2019 (COVID-19) pandemic in Lebanon along time. We have developed two different models of increasing complexity using Bayesian MCMC simulation methods, and found that the Poisson autoregressive as a function of both a short-term dependence and a long-term dependence provides the best fit to the data. The use of Poisson autoregressive model that allows to capture short and long term memory effects can greatly improve the estimation of number of new cases and can indicate whether disease has an upward/downward trend, and where about every country is on that trend, all of which can help the public decision-makers to better plan health policy interventions and take the appropriate actions to contain the spreading of the virus to the degree possible. The mode is applicable to other countries and more time periods as data becomes available.

## Data Availability

Data are available from the website of the Ministry of Public Health in Lebanon and worldometer website.

https://maps.moph.gov.lb/portal/apps/opsdashboard/index.html#/d19be998323548278e088076d46d24f8

